# HIV positive patients on HAART continue to have a decline in renal function irrespective of Tenofovir usage: A 2-year cohort from an Indian tertiary care centre

**DOI:** 10.1101/2022.09.30.22280576

**Authors:** Kavita S. Joshi, Viplove F. Jadhao, Rushabh Y. Gujarathi, Widhi Churiwala, Anuya A. Natu

**Affiliations:** Department of General Medicine, Seth GS Medical College and K.E.M Hospital, Mumbai; Seven Hills DCH, Mumbai; Seth GS Medical College and K.E.M Hospital, Mumbai

**Keywords:** HAART, Tenofovir, renal dysfunction, eGFR, Creatinine Clearance

## Abstract

**Context:** HIV patients may undergo renal damage due to disease or nephrotoxic drugs. Tenofovir has been associated with the development of renal impairment.

**Aims:** To study and compare trends in Creatinine Clearance (CrCl) and estimated Glomerular Filtration Rate (eGFR) in patients on Highly Active Antiretroviral Therapy (HAART) and to compare the same between patients on Tenofovir and non-Tenofovir based regimens.

**Methods:** A cross-sectional study was conducted. We followed 244 patients for a period of 2 years. The demographic, clinical, and laboratory parameters of the patients were recorded at baseline, one year of therapy, and two years of therapy. The data was analyzed by dividing patients into Tenofovir and non-Tenofovir based groups. Statistical analysis used the Chi-square test, paired and unpaired t-tests, and Fischer’s exact test.

**Results:** The mean BUN and serum creatinine in both groups were comparable at the start of the therapy. The decline in CrCl and eGFR in all patients on HAART for two years was statistically significant, irrespective of Tenofovir usage. The mean fall in eGFR in the Tenofovir group was 12.4 mL/min/1.73 m^2^ and in the non-Tenofovir group, 9 mL/min/1.73 m^2^, though the differences between eGFR and CrCl were not significant between the two groups at any point.

**Conclusions:** Even though HAART usage has been said to slow the decline in kidney function in PLHIV, patients who receive HAART still show a statistically significant decline in renal function parameters, akin to the observations of other such studies in low-resource settings.

## Introduction

Globally, there was an estimated prevalence of 37.7 million (30.2 – 45.1 million) people living with HIV at the end of 2020.^1^ India is considered a low prevalence country in terms of HIV (Human Immunodeficiency Virus), with an estimated adult prevalence of 0.22% and 23.49 lakh PLHIV (People Living with HIV/AIDS). Maharashtra reported a PLHIV prevalence, higher than the national average, of 0.36% and an estimated numerical burden of 3.96 lakh.^2^

Renal dysfunction in PLHIV is a widely studied topic, with various factors known to be contributing to the decline in renal function seen in such individuals. Along with an increased risk of both acute and chronic kidney disease, the mortality associated with renal disease is also said to be higher in PLHIV.^3^ The past few decades have seen a decline in mortality due to infectious complications from HIV worldwide, and subsequently, other HIV-related non-communicable diseases such as renal, cardio-metabolic, and liver disease have become increasingly common causes of mortality.^4^

The etiology of renal dysfunction in PLHIV is multifocal, including but not limited to HIVAN (HIV associated nephropathy), immune complex kidney disease, and drug-induced nephrotoxicity.^5^ An increased incidence of HIVAN was seen with lower CD4 counts, hence, a possible association with treatment failure may exist.^6^ Although the incidence of HIVAN has reduced with the widespread use of HAART, the nephrotoxic effects of HAART drugs is an important concern in the long-term management of HIV.^7^

As per the Sankalak Report and its previous editions by the National AIDS Control Organisation, a widespread rollout of Dolutegravir-based regimens was initiated in the country in recent years.^2^ One of the most frequently prescribed Dolutegravir-based regimens, known as the TLD regimen, contains Tenofovir (Tenofovir Disoproxil Fumarate 300 mg) + Lamivudine (3TC 300 mg) + Dolutegravir (DTG 50 mg) as a fixed drug combination in a single pill, usually given once a day.^8^

Tenofovir has been widely discussed as being associated with a low but significant risk of kidney injury.^9^ Although the study of safer alternatives is continually underway, the current widespread use of Tenofovir in Indian HAART regimens, along with the importance of renal disease as a cause of morbidity and mortality in PLHIV, makes it important to study various clinical implications of Tenofovir usage in this population, including but not limited to renal function.

### Rationale

Along these lines, we undertook this study to assess the creatinine clearance (CrCl) and the estimated glomerular filtration rate (eGFR) in HIV patients on various HAART regimens. The study was done to observe the trends of eGFR and CrCl at a Mumbai-based tertiary care centre and to describe the associations found within these trends.

## Methods

### Study Type

A prospective observational study was conducted at the virology/HIV outpatient department (OPD) and General Medicine wards of our institute, a Mumbai-based tertiary care centre.

### Study Population and Sample Size

Patients attending the OPD and admitted to the wards were recruited for the study and followed for a period of two years.

An approximate sample size was calculated using a formula proposed by Casagrande et al.^10^ A renal involvement prevalence in PLHIV of 17.3% was used based on a North Indian study by Gupta et al.^11^ The size of the population was approximated based on the number of PLHIV visiting the Virology OPD at our centre. Based on these estimates, the calculation yielded an estimated sample size of 250.

244 participants were enrolled in the study.

### Selection of Participants

The inclusion criteria involved all patients above 18 years of age who had been receiving HAART for a minimum period of 6 months. Patients with current urinary tract infections, those receiving known nephrotoxic drugs, those with Hepatitis B or Hepatitis C infections, or those known to have chronic kidney disease were excluded from the study. Following that, study participants were divided into two groups based on whether they received Tenofovir as part of their ART regimens:

1. Patients on Tenofovir based HAART
2. Patients on non-Tenofovir based HAART

### Methods of Measurement

A detailed history, including various demographic parameters, and physical examination were documented for all participants. A baseline biochemical data profile (mainly serum creatinine) and a baseline CD4 count were recorded for each patient. Subsequent data was collected from fresh blood samples taken at their follow-up visits after 1 year and 2 years from their date of enrolment, and the same biochemical parameters were recorded at both of these visits.

### Outcomes and Assessment

The primary outcomes measured were estimated glomerular filtration rate (eGFR) and Creatinine Clearance (CrCl). eGFR was calculated using the Chronic Kidney Disease Epidemiological Collaboration Equation (CKD-EPI).^12^ CrCl was calculated by using the Cockcroft-Gault equation. ^13^ These parameters were recorded at the time of enrolment, after one year of therapy, and after two years of therapy. Both groups were compared amongst themselves for the changes in all measured parameters at baseline, after 1 year, and after 2 years from the baseline. The groups were also compared with each other at these three intervals.

### Ethical Approval and Informed Consent

Institutional ethics committee and National AIDS Control Organization approvals were taken prior to the commencement of the study. Written informed consent was obtained from each patient. No changes were made to the ART regimens of the participants for the purpose of this study. Patients were recruited based on their existing treatment regimens.

#### Data Analysis/Statistics

Descriptive statistical analysis of data was done. Continuous data were categorized and the two study groups were compared using the Chi-square test or the Fischer’s exact test as applicable. The t-test (paired or unpaired as applicable) was used for analyzing the differences among the means of quantitative parameters. A P value of 0.05 was used across all as a cut-off for significance. All statistical analysis was done using SPSS Version 28.^14^

## Results

A total of 244 patients were enrolled in the study, of whom 123 were on Tenofovir-based therapy (Tenofovir as the main component of their therapy) and 121 were on non-Tenofovir-based therapy.

In the Tenofovir-based therapy (*N = 123*) group, most patients were on Tenofovir, Lamivudine, and Efavirenz (TLE) (*N = 118, 95*.*93%*), while five patients (*N = 5, 4*.*07%*) were on Tenofovir, Lamivudine, and Nevirapine (TLN). In the non-Tenofovir-based therapy group (*N = 121*), the highest number of patients were on Zidovudine, Lamivudine, and Nevirapine (ZLN) (*N = 88, 72*.*73%*), followed by Stavudine, Lamivudine, and Nevirapine (SLN) (*N = 12, 9*.*92%*), Zidovudine, Lamivudine, and Efavirenz (ZLE) (*N = 11, 9*.*09%*), and Stavudine, Lamivudine, and Efavirenz (SLE) (*N = 10, 8*.*26%*).

At baseline, there were no significant differences between the two groups in their demographic parameters, clinical exam findings, past medical history/comorbidities, or biochemical investigation results.

Upon comparing the mean eGFR values between the two groups, the Tenofovir group had a higher mean eGFR value to begin with, but there was a greater decline in eGFR in the Tenofovir group as compared to the non-tenofovir group. This difference, however, was not found to be statistically significant at any of the three points of measurement.

The mean CrCl values between the two groups followed a similar trend in terms of overall decline in both groups. In contrast to eGFR, the values remained higher in the Tenofovir group at all three points of measurement. However, like the eGFR trends, none of the differences were found to be statistically significant.

Using the paired t-test, CrCl and eGFR values were compared across the entire cohort for any differences in values between baseline and 1-year and 2-year post-baseline measurements. There was a statistically significant difference in both eGFR and CrCl values when comparing the baseline with the values at both 1- and 2-years post-baseline. There was a significant difference between the values at the 1-year and 2-year marks as well for both the parameters. The difference between the mean reduction in eGFR or CrCl was not significant between the two groups.

The distribution of patients across various stages of kidney disease based on eGFR, as described by the KDIGO is given in Table IV.^15^

The mean CD4 counts were found to rise across the whole cohort throughout the entire study period. The non-tenofovir group showed a greater rise. The mean CD4 counts in the Tenofovir group went from 484.7 at baseline to 530 at 1-year of measurement post-baseline, followed by 539 at the 2-year mark. The mean CD4 counts in the non-tenofovir group rose from 537 at baseline to 555 and 614 at the 1-year and 2-year post-baseline marks, respectively.

## Discussion

Due to our inclusion criteria, most patients with hypertension and diabetes mellitus were excluded from the study population, and both the therapy groups were comparable, as demonstrated in Table I. This gave our study a unique perspective on renal function in PLHIV, as subjects were studied without the presence of two of the most common comorbidities that are associated with renal impairment worldwide.^16^ As a result, we were able to focus the analysis on the HAART regimens received by the participants, which was in line with the study’s goals.

**Table I:**
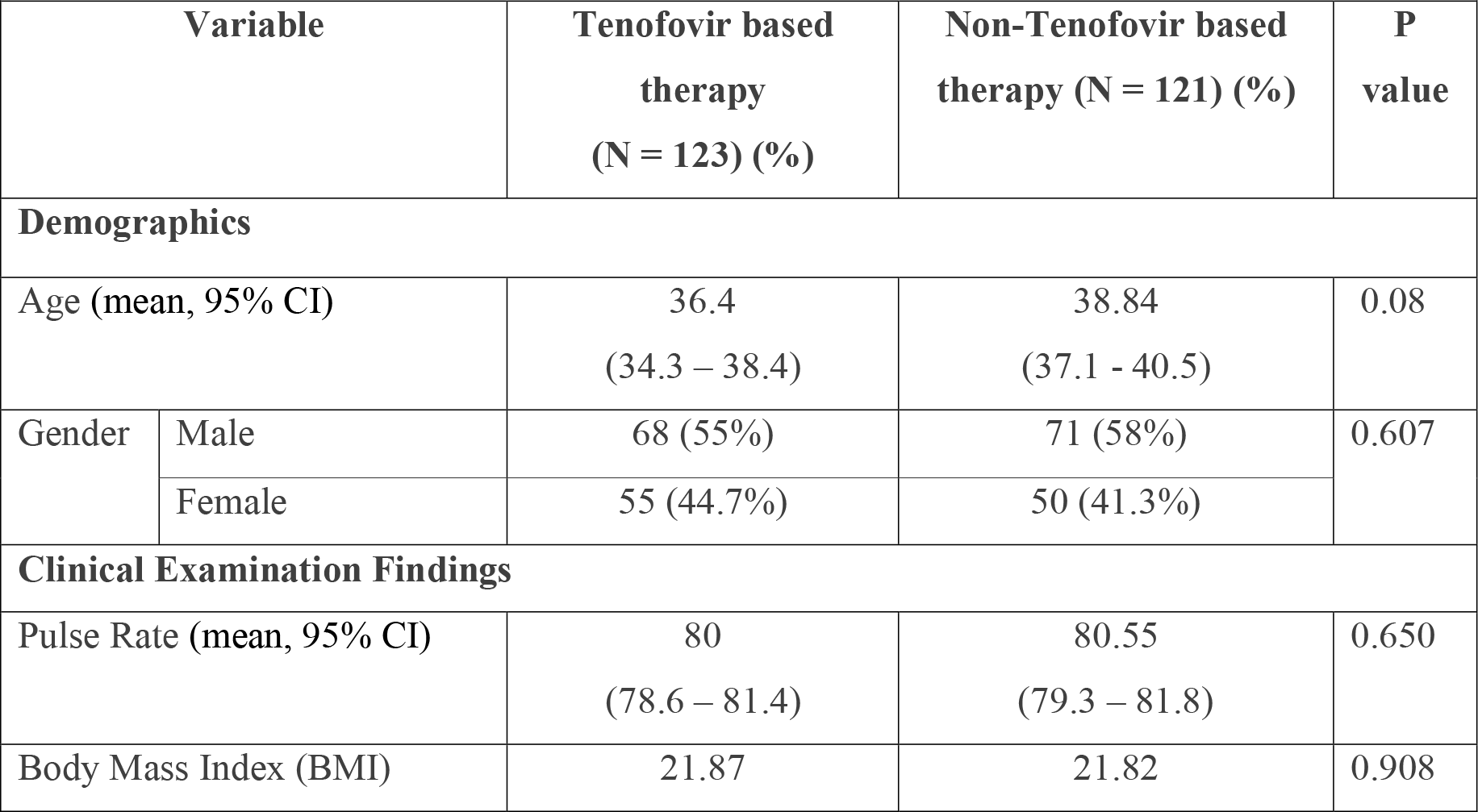
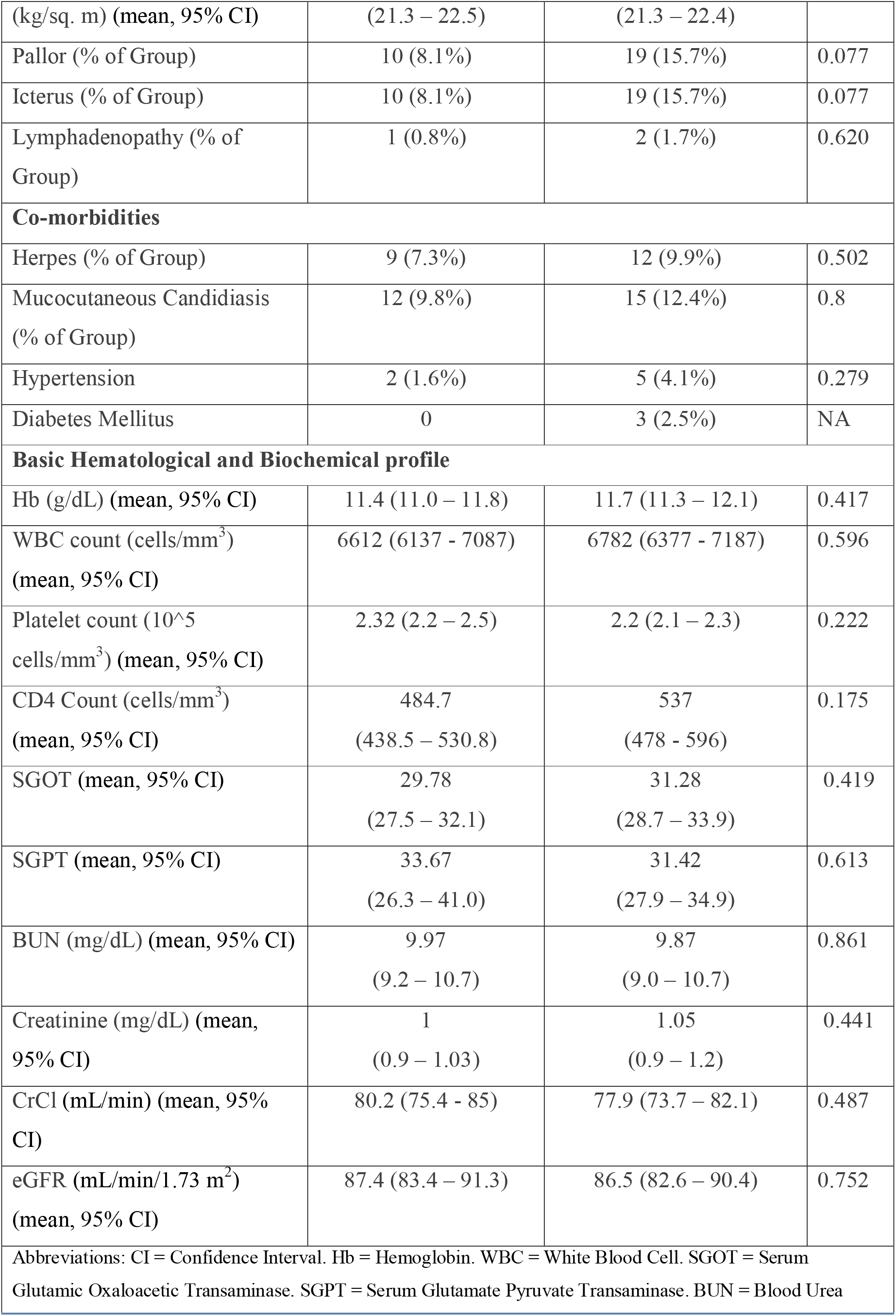
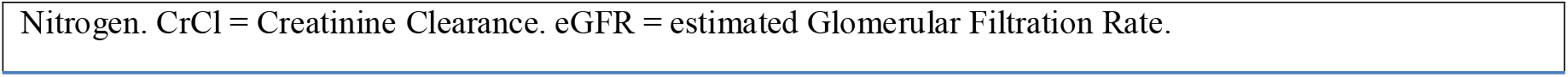
Comparison of select parameters and findings between the two groups at baseline.

**Table II:** Trends in eGFR and CrCl across 2 years in the entire cohort.

| **Trends in eGFR** |  |  |  |  |
| --- | --- | --- | --- | --- |
| Sr. No. | Interval of Measurement | eGFR in Tenofovir based group (mL/min/1.73 m<sup>2</sup>) | eGFR in non-tenofovir based group (mL/min/1.73 m<sup>2</sup>) | P value |
| 1. | Baseline (mean, 95% CI) | 87.4 (83.5 – 91.3) | 86.50 (82.6 – 90.4) | 0.752 |
| 2. | At 1 year after the baseline (mean, 95% CI) | 82.7 (78.7 – 86.6) | 83.8 (79.9 – 87.7) | 0.700 |
| 3. | At 2 years after the baseline (mean, 95% CI) | 75 (71 – 78.9) | 77.5 (73.6 – 81.4) | 0.382 |
| **Trends in CrCl** |  |  |  |  |
| Sr. No. | Interval of Measurement | CrCl in Tenofovir based group (mL/min) | CrCl in non-tenofovir based group (mL/min) | P value |
| 1. | Baseline (mean, 95% CI) | 80.2 (75.4 – 84.9) | 77.9 (73.7 – 82.1) | 0.487 |
| 2. | 1 year after the baseline measurement (mean, 95% CI) | 75.4 (70.6 – 80.2) | 74.9 (70.6 – 79.1) | 0.879 |
| 3. | 2 years after the baseline measurement (mean, 95% CI) | 71.6 (66.8 – 76.4) | 70.6 (66.3 – 74.8) | 0.761 |
| Abbreviations: CI = Confidence Interval. CrCl = Creatinine Clearance. eGFR = estimated Glomerular Filtration Rate. |  |  |  |  |

A study by Gallant et al. showed that patients with a lower CD4 cell count, decreased renal function at baseline, and diabetes mellitus show a modest decline in creatinine and creatinine clearance during the study period. In the same study, the most evident feature was that the use of Tenofovir Disoproxil Fumarate (TDF) was associated with a greater decline in renal function (defined as a change in creatinine clearance) than the use of alternative nucleoside analogues. Although statistically significant, the decline in CrCl associated with TDF use was small and was not associated with a greater rate of discontinuation.^17^ Along similar lines, patients at our center did not require changes in their HAART regimens despite a statistically significant decline in CrCl in both groups after two years.

The most important observation in our study was the decline in eGFR and CrCl despite the use of HAART. Calle et al. described a significant decline in renal function in a long term observational prospective cohort study studied over 7.5 years and attributed this change to the use of Tenofovir.^18^ Kumarasamy et al. described a similar relationship with Tenofovir use.^19^ Mulenga et al. also found in their adjusted analysis that patients receiving TDF were more likely to experience an episode of moderate or severe renal dysfunction than those receiving other regimens during the first year of ART, although the incidence of such episodes was low.^20^

As shown in Table IV, we observed that a total of 168 patients showed a fall in eGFR out of the 244 study participants. We observed that 6 patients showed a decline in eGFR of more than 60 mL/min/1.73 m^2^ in two years from the baseline, out of which 5 patients were from the Tenofovir-based therapy group and 1 was from the non-Tenofovir group. Although not significant, this difference shows a possible need for longer cohorts analyzing Tenofovir use in urban Indian populations such as ours.

**Table III:** Temporal analysis of CrCl and eGFR values using the paired t-test across the whole cohort.

|  |  |  |  |
| --- | --- | --- | --- |
| Sr. No | Comparison between | t | P-value |
| **Creatinine Clearance comparisons** |  |  |  |
| 1 | CrCl at baseline vs. CrCl at 1 1-year post-baseline | 2.937 | 0.004 |
| 2 | CrCl at baseline vs. CrCl at 2-years post-baseline | 5.726 | < 0.001 |
| 3 | CrCl at 1-year post-baseline: CrCl at 2-years post-baseline | 4.996 | < 0.001 |
| **eGFR comparisons** |  |  |  |
| 1 | eGFR at baseline: eGFR at 1-year post-baseline | 2.85 | 0.005 |
| 2 | eGFR at baseline: eGFR at 2 years | 7.037 | < 0.001 |
| 3 | eGFR at 1-year post-baseline vs. eGFR at 2 years | 5.677 | < 0.001 |
| Abbreviations: CrCl = Creatinine Clearance. eGFR = estimated Glomerular Filtration Rate. |  |  |  |

**Table IV:**
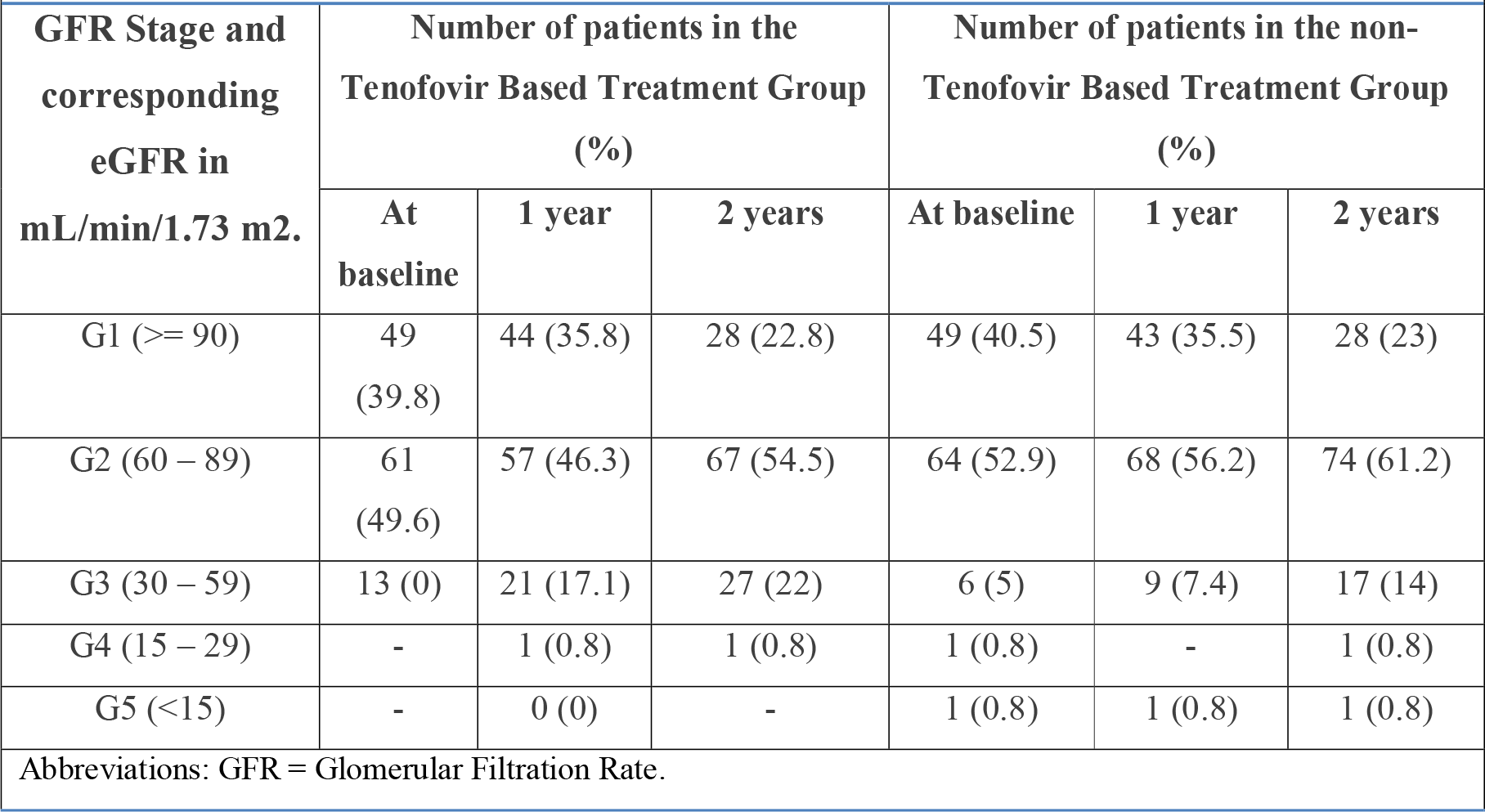
Distribution of study subjects based on stages of kidney disease.

A retrospective cohort analysis of 1,647 HIV-infected patients by Michael Horberg et al. defined renal dysfunction as changes in glomerular filtration rate and reported Tenofovir usage as having a greater impact on decline in renal function. They also reported an increased incidence of proximal tubular dysfunction associated with Tenofovir usage.^21^ Our study, however, did not show any statistically significant difference in the CrCl and eGFR values of patients between the Tenofovir and non-Tenofovir groups. A smaller sample size in our study and the exclusion of patients with possible effect modifiers like diabetes mellitus and hypertension might be a possible explanation for this lack of association in our study. Nayak et al. also found no statistically significant difference in creatinine clearance as well as eGFR after 6 months and 12 months of treatment when two groups treated with Tenofovir and drugs other than Tenofovir were compared.^22^

An increase in the average CD4 counts of our study population indicates that therapeutic failure may not be considered as a reason for the declines seen in our study. The similarity in CD4 counts across the two study groups (*P = 0*.*07*) shows that therapeutic responses might also be similar with both types of HAART regimens. Although Lucas et al. reported a decline in the incidence of HIVAN across a 12-year cohort with HAART usage, findings from other studies across various settings are a cause for concern.^6^ A 6-year cohort by Fiseha et al. reported a decline in renal function in patients being started on HAART in Ethiopia.^23^ Szczech et al. also reported an absence of any beneficial effect of HAART or viral suppression on the decline in renal function observed in their study.^24^

Pujari et al., through a study conducted at both the Institute of Infectious Diseases (IID) Pune, Western India, and the Royal Free Hospital (RFH) London, observed that there was a statistically significant decline in eGFR from baseline at 6 and 24 months at both the sites. However, there was no statistically significant difference in eGFR decline between the two sites. People living with HIV at both sites had an average decline in eGFR of 7 mL/min/1.73 m^2^ at 24 months post-TDF initiation.^25^ The results observed in our study resembled these findings, with the tenofovir group showing a significant decline in mean eGFR of 12.4 mL/min/1.73 m^2^. Notably, the non-Tenofovir group also demonstrated a similar (significant) decline of 9 mL/min/1.73 m^2^ in mean eGFR in our study.

The intention of the present study was not limited to analyzing Tenofovir as a direct cause of renal function decline in PLHIV. We aimed to look for renal dysfunction in PLHIV at an urban Indian tertiary care center and look for risk factors (demographic, clinical, biochemical, disease progression, and therapy-related) associated with renal impairment in PLHIV receiving HAART. The exclusion of most diabetics and hypertensives based on our inclusion criteria presents a data set which possibly reflects the effects of other factors influencing renal function. Although similar studies have shown an association between tenofovir usage and the decline of renal function in PLHIV, we believe that the findings from our study and similar others, which show an overall decline irrespective of ART regimen, should not be ignored. A single drug or therapy regimen cannot be attributed to renal dysfunction in this population. In the face of potential widespread usage of Tenofovir through the DLT regimen in India, further studies assessing renal function and careful monitoring of renal disease are suggested.

### Limitations

Our study had a small sample size and a short follow-up period. Since this was an observational study, histopathological samples were not obtained, and hence we might have missed possible diagnostic and etiological clues, which would have been more evident with renal biopsies.

## Data Availability

All data produced in the present work are contained in the manuscript

## Conflicts of Interest

Nil.

## Notes

Source(s) of financial support: nil.

Conflicts of Interest: none – for all the authors.

### Competing Interest Statement

The authors have declared no competing interest.

### Funding Statement

This study did not receive any funding

### Author Declarations

Ethics Committee of Seth GS Medical College gave ethical approval for this work

